# Lived Experiences of Never-Married Aging Nurses in Clinical Practice: A Qualitative Study

**DOI:** 10.64898/2026.03.02.26347293

**Authors:** Fernan Torreno, Frincess Flores

## Abstract

The global nursing workforce is aging, yet limited research has explored the lived experiences of never married nurses entering midlife and later adulthood. Existing studies have primarily focused on burnout and retention, with less attention to the social and existential dimensions of aging without a spouse or children. This study aimed to explore the experiences of never married clinical nurses aged 40 years and older, focusing on perceptions of aging, professional identity, social support, and future security.

A qualitative descriptive design was employed. Twenty-five never married nurses aged 44–62 years were recruited through purposive sampling from intensive care, emergency, medical, surgical, oncology, outpatient, and community departments across four government hospitals. Semi-structured interviews were conducted and analyzed using reflexive thematic analysis. Trustworthiness was ensured through member checking, peer debriefing, and maintenance of an audit trail.

Four themes were identified: **Nursing as a Life Anchor**, where professional identity provided meaning and structure; **Independence Coexisting with Loneliness**, reflecting autonomy alongside episodic loneliness; **The Invisible but Available Workforce**, describing expectations of greater work availability due to single status; and **Anticipating an Uncertain Future**, capturing concerns about retirement, declining health, and limited advocacy in later life.

Never married aging nurses experience a complex balance of professional fulfillment, autonomy, vulnerability, and uncertainty. Healthcare organizations should recognize this subgroup and consider equitable workload policies, tailored retirement planning, and psychosocial support to promote well-being and workforce sustainability.

## Introduction

Although increasing numbers of students are entering nursing programs, the global nursing workforce is undergoing significant demographic change as large numbers of nurses approach retirement age. Projected worldwide shortages are expected to worsen over the next decade due to population aging and the retirement of experienced nurses^1,2^. The World Health Organization estimates that the global shortage of nurses may reach several million by 2030 unless retention efforts are strengthened^1^. The International Council of Nurses has identified workforce aging as a crisis that poses a serious threat to the sustainability of health systems^2^.

Older nurses contribute substantial clinical expertise, institutional knowledge, and mentorship to healthcare organizations. They frequently serve as role models and preceptors for younger colleagues. As medical complexity increases and populations age, the need for experienced nurses remains critical. However, aging is accompanied by physiological changes that may affect physical capacity and endurance, including reduced strength, decreased mobility, fatigue, and chronic health conditions. Research on work ability indicates that aging workers benefit from supportive workplace modifications, such as ergonomic adjustments and flexible scheduling, to maintain productivity and well-being_. Nursing is both physically and emotionally demanding, involving irregular hours, rotating shifts, prolonged standing, and exposure to patient suffering. These demands may intensify the challenges associated with aging.

Retention of the aging nursing workforce has therefore become a global priority. Studies indicate that supportive leadership, professional recognition, and positive work environments contribute to extended career participation among older nurses_,_. In contrast, heavy workloads, inadequate staffing, and limited organizational support are associated with burnout and early retirement intentions_. The COVID-19 pandemic further underscored both the resilience and vulnerability of older nurses and highlighted the need for equitable workplace policies^1^_.

While workforce research has extensively examined burnout, turnover intention, and job satisfaction, relatively little attention has been paid to how personal life circumstances intersect with aging and professional identity. Never-married nurses represent a subgroup whose experiences may differ from those of married colleagues with established family support systems. Gerontological literature consistently demonstrates that social support is a major determinant of psychological well-being, health outcomes, and successful aging^11,12^. Individuals aging without close familial support may experience heightened concerns regarding loneliness, future care, and personal vulnerability^13^.

Assumptions regarding family responsibilities may also influence workplace expectations. Qualitative studies suggest that nurses without spouses or children are often perceived as more available for overtime or shift coverage^1^_. Such expectations may inadvertently create inequities in workload distribution. For aging nurses, this dynamic may be compounded by age-related changes in physical recovery capacity_. Despite these realities, there remains limited research specifically focused on the experiences of never-married aging nurses in clinical practice.

Professional identity is another important dimension of nurses’ well-being and career longevity. A strong professional self-concept is positively associated with work engagement and negatively associated with turnover intention^1^_. For many aging nurses, identity as a caregiver and experienced practitioner serves as both a coping resource and a central life anchor. However, the relationship between professional identity and living alone has not been adequately explored in nursing research.

Life course theory emphasizes that aging is shaped by the cumulative effects of work experiences, social relationships, and broader societal contexts^1^_. For individuals who have remained unmarried, the experience of aging may be shaped by different social trajectories compared with those who followed more traditional family pathways. Research suggests that individuals living alone in later life may have increased risk of perceived social isolation and health-related anxiety compared to those living with partners^1^_, although greater autonomy and independence are also frequently reported^1^_. Understanding how these dynamics manifest within the nursing profession is important for workforce planning and supportive policy development.

Retirement planning presents additional considerations for single individuals. While financial preparation is essential, emotional security and future caregiving arrangements are equally significant. Nurses, because of their professional exposure to patient vulnerability and end-of-life care, may possess heightened awareness of these concerns. Nevertheless, few qualitative studies have examined how aging nurses conceptualize their own future support needs.

Age-friendly work environments that value experience and accommodate physical and psychosocial changes are essential for sustaining the aging workforce^2^,_. However, many institutional policies implicitly assume the presence of family support structures. Consequently, the needs of never-married aging nurses may remain unrecognized in workforce planning.

Qualitative inquiry provides an opportunity to explore these underexamined dimensions. By examining lived experiences, researchers can better understand how never-married aging nurses interpret independence, loneliness, professional identity, workload expectations, and future planning within their sociocultural context^1^_.

Given the global aging of the nursing workforce and the paucity of research addressing relationship status as a contextual factor, this study aimed to explore the lived experiences of never-married nurses aged 40 years and older in clinical practice. By centering their narratives, this study seeks to contribute to a more inclusive understanding of aging within the nursing profession and to inform equitable workforce policies.

## Methods

### Study Design

Although never-married aging nurses constitute a growing sector of the global nursing workforce, there is a paucity of empirical research focused on their unique experiences. This study employed a qualitative descriptive design to explore the lived experiences of never-married aging nurses. A qualitative descriptive approach allows for detailed exploration of the phenomenon of interest while remaining close to the language and meaning of participants. It provides a contextualized description of the experiences of never-married aging nurses and how they perceive aging, the nursing profession, and future planning within clinical settings.

### Setting and Participants

The study took place in Region I of the Philippines across three types of healthcare facilities, including tertiary hospitals, secondary hospitals, and community health facilities. The clinical areas involved included intensive care units, emergency departments, medical and surgical wards, oncology units, outpatient departments, and community health services.

Purposive sampling was employed from a defined population of registered nurses who met the inclusion criteria. To ensure variability in age, years of practice, and clinical discipline, registered nurses aged 40 years and older who were never married, were not living with a partner or spouse, had a minimum of five years of clinical experience, and were actively practicing in a clinical capacity were recruited for the study. Divorced and widowed nurses and those cohabiting with a partner were excluded to maintain conceptual homogeneity of the “never-married” subgroup.

Recruitment continued until data saturation was achieved. Saturation was defined as the point at which no new themes and no meaningful new information emerged from subsequent interviews. Saturation was observed after twenty-three interviews; two additional interviews were completed to verify that the themes had stabilized. A total of twenty-five participants took part in the study.

### Data Collection

Data were collected between [January to February 12, 2026] via semi-structured, in-depth interviews. Interviews took place in private meeting rooms at participants’ worksites or through a secure online video platform, depending on participant preference and feasibility. Interviews lasted approximately one hour to one and a half hours.

The interview guide was developed following a review of relevant literature and in consultation with qualitative researchers. Questions explored participants’ subjective experiences of aging, professional identity, perceptions of workplace expectations, social networks, and experiences of loneliness or autonomy. Participants were also asked about views on retirement and long-term economic security. Questions were primarily open-ended to elicit narrative responses, and probing questions were used to clarify meaning and encourage elaboration.

All interviews were audio-recorded with informed consent and transcribed verbatim. Transcripts were anonymized by assigning participant codes from P01 to P25. Field notes were completed soon after each interview to document contextual details and researcher reflections.

### Data Analysis

Data analysis was conducted using reflexive thematic analysis. The research team began by reading transcripts multiple times to achieve immersion in the data. Initial codes were developed through line-by-line coding, focusing on meaningful segments. Codes were compared across transcripts to identify patterns and relationships, and then organized into themes and subthemes. Themes and subthemes were refined through iterative discussion among the researchers.

NVivo software (version XX) was used to organize and manage the data. Analytic decisions were documented throughout the process to ensure transparency. Differences in coding and interpretation were discussed during team meetings until agreement was reached.

### Rigor

To ensure rigor, strategies were applied to support credibility, dependability, confirmability, and transferability. Credibility was strengthened through member checking with five participants who reviewed summaries of preliminary themes. Dependability was supported through maintenance of an audit trail documenting methodological and analytic decisions. Confirmability was enhanced through reflexive journaling, including documentation of researcher assumptions and positionality. Transferability was supported by providing a clear description of participants and research context to enable readers to assess applicability to other settings.

### Ethical Considerations

Ethical approval was obtained from the Philippine Region I Ethical Review Committee (Approval No. NUR-2026-3076A). All participants provided written informed consent prior to data collection. Informed consent was conducted as a two-way discussion in which the researcher explained the voluntary nature of participation and the right to withdraw at any time without penalty.

Participant confidentiality was protected by removing all personally identifiable information from transcripts. All electronic data were stored securely using password-protected devices and files accessible only to the research team. Given the sensitive nature of discussing personal experiences, participants were informed about the availability of psychosocial support services should distress arise. No participants requested referral for psychosocial suppor

## Results

The study involved twenty-five never-married nurses aged 44 to 62 years. Participants worked in a range of clinical settings including intensive care, emergency, medical–surgical wards, oncology, outpatient clinics, and community health. All participants had between 19 and 38 years of professional nursing experience. The majority lived alone, while a small minority lived with elderly parents or provided a caregiving role for aging parents. The demographic characteristics of the participants are presented in Table 1.

**Table 1.**
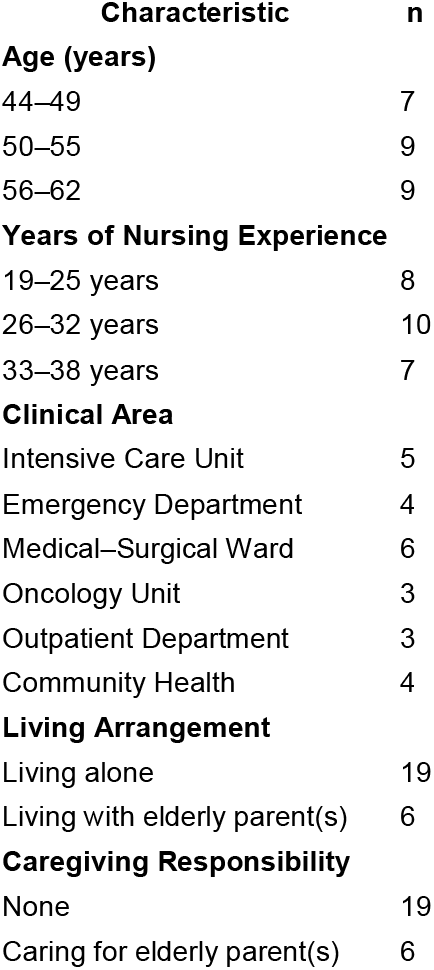
Demographic Characteristics of Participants (N = 25)

Four main themes emerged from the interviews: Nursing as a Life Anchor, Independence Coexisting with Loneliness, The Invisible but Available Workforce, and Anticipating an Uncertain Future. These themes were multidimensional and integrated multiple aspects of aging, professional identity, social context, and their interconnections. The themes and corresponding subthemes identified through reflexive thematic analysis are summarized in Table 2.

**Table 2.** Themes and Subthemes Identified Through Reflexive Thematic Analysis.

| Theme | Subthemes |
| --- | --- |
| **Nursing as a Life Anchor** | Professional identity as stability and meaning<br>Clinical expertise and mentorship<br>Age-related recognition and perceived limitations |
| **Independence Coexisting with Loneliness** | Financial and emotional self-reliance<br>Episodic and situational loneliness<br>Navigating family-centered social interactions |
| **The Invisible but Available Workforce** | Assumed scheduling flexibility<br>Perceived inequity in workload distribution<br>Limited institutional recognition of aging alone |
| **Anticipating an Uncertain Future** | Concerns about illness and advocacy<br>Retirement and loss of routine<br>Financial preparedness versus emotional security |

### Nursing as a Life Anchor

Nursing was described as stable and enduring and as a central factor providing meaning and structure in participants’ lives. Many participants described nursing as their main life commitment.

> “Nursing has been the most constant relationship in my life” (P12).

Participants described work not only as employment but also as a role and identity. They expressed pride in clinical skills and in the coping abilities developed through decades of experience. Participants reported that age brought wisdom, patience, and greater clinical judgment even as physical strength and endurance decreased.

> “I am more tired physically now, but I am calmer and more confident in my decisions” (P13).

Recognition from younger colleagues contributed to participants’ sense of value and reinforced professional identity. However, participants also described perceived age-related barriers to progression or leadership opportunities.

> “After fifty, you are seen as experienced but not necessarily promotable” (P17).

Some participants reflected on broader institutional tensions affecting practice environments, including perceived gaps between service expectations and training systems, while also acknowledging the role of professional development pathways in creating structure, learning opportunities, and social connection within nurses’ daily lives.

### Independence Coexisting with Loneliness

Independence was frequently emphasized when participants described being never married. Participants described themselves as independent decision-makers and financially self-reliant, often framing these qualities as strengths that reduced emotional vulnerability.

> “I learned to rely on myself. That is my strength” (P20).

For some participants, never being married was viewed positively because it allowed flexibility in personal schedules and the freedom to pursue individual interests. At the same time, many participants described episodic loneliness, particularly during holidays and after demanding shifts.

> “After the night shift, my home is silent. That is when I feel it most” (P01).

Participants described how social interactions could be shaped by family-centered conversations, which sometimes led them to feel excluded. One participant noted that conversations frequently focused on children and grandchildren and that she occasionally felt outside those patterns (P07).

Participants emphasized that loneliness was not constant but situational and episodic. Many described coping by developing friendships, engaging in leisure activities, focusing on personal well-being, and maintaining social ties. Across accounts, participants conveyed the emotional complexity of balancing independence with intermittent experiences of loneliness.

### The Invisible but Available Workforce

A common thread across interviews related to workload expectations and scheduling. Many participants perceived that because they were single, others assumed they were more available for overtime, shift changes, or additional responsibilities.

> “If someone needs to swap shifts for family reasons, they usually ask me” (P15).

Although participants expressed empathy for colleagues with family caregiving responsibilities, they also felt their own rest and personal time could be undervalued.

> “Just because I am single does not mean my rest is less important” (P22).

Participants described feeling “invisible but available,” referring not only to scheduling but also to broader experiences of being overlooked by workplace policies that they perceived as primarily designed around family-related needs. Participants felt that institutional supports often prioritized childcare or parenting needs without similarly addressing concerns related to aging alone.

> “There are programs for parents, but none really for someone aging alone” (P11).

Although participants identified these dynamics as problematic, many described them as normalized within workplace culture and tended to avoid directly challenging them.

### Anticipating an Uncertain Future

Concerns about aging and retirement were frequently raised. While many participants reported awareness of financial planning and saving for the future, they expressed less certainty about emotional and social readiness for later life.

> “Money can be saved, but companionship cannot be planned” (P02).

Participants frequently discussed worries about illness and the potential lack of advocacy during healthcare encounters.

> “I worry about who will speak for me if I cannot speak for myself” (P03).

Participants expressed ambivalence about retirement. Some anticipated retirement as rest after long service, while others feared loss of routine and reduced social connection.

> “When I stop working, I lose my daily connection to people” (P16).

In earlier interviews, participants’ narratives often emphasized fear and uncertainty regarding later life. As discussion continued, participants also described coping strategies and proactive planning, including maintaining health, strengthening friendships, and preparing practical arrangements for the future. These accounts reflected adaptation and acknowledgment of changing roles, alongside a desire for more organized institutional recognition of their circumstances.

Overall, participants described their working and personal lives across four intersecting dimensions: contentment and meaning derived from professional identity, experiences of independence and episodic loneliness, perceived inequities in workload expectations, and anxiety about future health and retirement. These dimensions demonstrated variability across participants and highlighted both resilience and unmet support needs among never-married aging nurses.

## Discussion

This qualitative study explored the lived experiences of never-married aging nurses in Region I, Philippines. The findings showed that participants’ experiences were closely interconnected with professional identity, independence, workplace norms, and future-oriented concerns. Participants’ narratives reflected both universal aspects of aging in nursing and experiences shaped by social and cultural expectations.

A major theme was Nursing as a Life Anchor. Participants described nursing as the most enduring aspect of their lives, providing stability, meaning, and structure. This aligns with international literature suggesting that professional identity acts as a coping resource and supports continued workforce participation among older nurses. Participants reported satisfaction in practicing clinical skills and supporting younger colleagues through informal mentoring and guidance. In the Philippine context, nursing is a respected profession and an important pathway for economic mobility, and long-term professional investment may take on greater significance for those who do not occupy traditional family roles. For never-married nurses, the workplace was not only a site of employment but also a primary source of social interaction and belonging.

Alongside autonomy, participants described Independence Coexisting with Loneliness. While financial self-reliance and personal freedom were sources of pride, episodic loneliness emerged, particularly during time away from work and during family-focused social occasions. Consistent with gerontological research, living alone was not equated with constant loneliness; rather, loneliness was described as situational and intensified during specific life moments. Participants also described feeling less visible in family-centered social interactions, which may reflect cultural primacy given to marriage and family within collectivist contexts. Their accounts suggest that being never married may shape experiences of belonging, identity, and social recognition in later life.

The theme The Invisible but Available Workforce highlighted a perceived workplace norm in which single nurses were assumed to be more available for additional duties. Participants described being asked to cover extra shifts or overtime and perceived that these expectations were connected to marital status rather than formal policy. This dynamic is relevant to workforce equity and retention, particularly for older nurses who may require more recovery time from physically demanding shifts. Such experiences may influence morale and could contribute to early retirement intentions if left unaddressed.

Future-oriented concerns were captured in Anticipating an Uncertain Future. Participants differentiated between financial security and emotional security. While many described proactive financial planning, they expressed worries about illness, advocacy, companionship, and long-term care arrangements. These concerns mirror those raised in broader literature on aging alone. In the Philippines, where family care is a common traditional approach to elder support, the absence of immediate family advocacy may intensify perceived vulnerability. Participants’ professional exposure to patient dependency and end-of-life care may further heighten awareness of these concerns.

Despite challenges, participants described resilience and coping strategies, including health maintenance, self-care, and building social networks. This aligns with research suggesting that older nurses have developed substantial coping capacity through years of clinical experience. However, resilience does not replace organizational responsibility. Many healthcare institutions prioritize family-friendly arrangements such as parental leave and childcare support, which are essential, but participants’ narratives indicate that aging alone may introduce different support needs that should also be considered in workforce planning.

The findings have implications for workforce sustainability and nursing leadership. Assumptions based on marital status should be removed from scheduling practices and workload allocation. Clear and transparent policies regarding overtime and shift distribution may reduce perceptions of inequity. Retirement planning initiatives should incorporate not only financial preparation but also psychosocial planning, long-term care planning, and support for developing social networks. Workplace-based supports, such as peer mentoring, wellness initiatives, and psychosocial programs targeted to midlife and older nurses, may strengthen connectedness and well-being.

This study contributes to an under-researched area by giving voice to never-married aging nurses and highlighting how marital status intersects with professional identity, workload expectations, social experience, and future planning. Rather than treating marital status as a neutral background characteristic, the findings draw attention to underlying assumptions regarding caregiving responsibilities and availability that may influence work experiences. By examining participants’ accounts, the study highlights diversity within the aging nursing workforce and the need for inclusive, age-responsive policy development.

Several limitations should be noted. The findings reflect one region of the Philippines and may not transfer directly to different cultural or institutional contexts. Participation was voluntary, and those who chose not to disclose personal experiences may have differed from those who participated. The qualitative approach provides in-depth insights into a specific context rather than generalizable estimates. Future research comparing never-married, married, and previously married aging nurses may further illuminate how relationship status interacts with work and aging. Quantitative research documenting workload distribution by marital status may also be valuable for policy development.

Ultimately, never-married aging nurses balance autonomy and vulnerability in everyday life. While their professional identity provides stability and meaning, concerns regarding future care requirements and workplace expectations remain salient. Recognizing this subgroup and integrating their needs into workforce planning is essential for developing age-responsive, culturally sensitive, and equitable nursing policy.

### Limitations

In considering study limitations, transferability of the findings is restricted to the context of Region I, Philippines. Future studies may explore whether similar findings occur across other regions of the Philippines and in other countries. Experiences of never-married aging nurses may differ in settings with different social values, healthcare systems, and institutional policies.

Second, purposive sampling and the voluntary nature of participation may have contributed to self-selection bias. Participating nurses may have been more reflective or more willing to discuss personal experiences, and some perspectives may not have been captured.

Third, self-reported interview data may be influenced by recall bias and social desirability bias, potentially understating negative workplace experiences or downplaying personal challenges.

Fourth, researcher interpretive bias is inherent in qualitative research. Although member checking and reflexivity were used to minimize bias, interpretation cannot be completely eliminated. Reflexive journaling and team discussion were used to strengthen confirmability, but complete objectivity is not possible.

Finally, the study focused specifically on never-married nurses. Direct comparison with married, divorced, or widowed nurses was outside the scope of this research. Future studies incorporating multiple relationship status categories may deepen understanding of how life circumstances intersect with aging and nursing practice.

## Conclusion

This study explored the lived experiences of never-married aging nurses in clinical settings and examined how professional identity, independence, and workplace circumstances intersected with future concerns. The findings revealed tensions related to professional identity, independence, workplace expectations, and uncertainty about later life. Nursing served as a life anchor, providing order, meaning, and social connection. Participants demonstrated resilience and independence developed through years of professional and personal self-reliance.

At the same time, work–life balance concerns, episodic loneliness, perceived inequities in workload expectations, and anxiety about retirement and long-term care were evident. Participants distinguished between financial and emotional security and expressed concerns about advocacy and companionship in later life. Recognizing the diversity of life circumstances within the aging nursing workforce is essential for equitable workforce planning and supportive policy development.

## Implications for Nursing Practice and Policy

The findings have implications for nursing administration, workforce planning, and hospital policy. Healthcare organizations should ensure that workload distribution and scheduling practices are not influenced by assumptions based on marital status. Clear scheduling guidelines and transparent approaches to overtime allocation may reduce perceptions of inequity.

There is also a need to develop retirement planning services that incorporate both financial preparation and psychosocial planning, including long-term care planning and strategies to strengthen support networks, recognizing that some nurses may lack immediate family-based support. Peer support and wellness initiatives geared toward midlife and older nurses may help strengthen social connections and mental well-being.

Recognizing never-married aging nurses as an identifiable subgroup within workforce planning may contribute to retention strategies. Policies and practices that acknowledge the contributions of experience while accommodating physical and psychosocial changes can enable older nurses to continue practicing clinically. Nursing leaders should consider diverse personal circumstances and avoid assuming the presence of supportive family structures. By acknowledging variability across life situations, leaders can provide equitable recognition and support for all staff.

## Declarations

### Ethics Approval and Consent to Participate

Ethical approval was obtained from the Philippine Region I Ethical Review Committee (Approval No. NUR-2026-3076A). All procedures were carried out in accordance with institutional ethical standards and the principles of the Declaration of Helsinki. Written informed consent was obtained from all participants. Participants were informed that participation was voluntary and that they could withdraw at any stage without penalty.

### Consent for Publication

All participants provided consent to be included in the study and consented to the use of anonymized quotations in publications arising from the research. All personally identifiable information has been omitted to protect confidentiality.

### Availability of Data and Materials

The qualitative dataset generated and analyzed during the current study is not publicly available due to the sensitive nature of the interview content and to protect participant confidentiality. De-identified excerpts may be made available from the corresponding author upon reasonable request and subject to ethical approval.

### Authors’ Contributions

All authors made substantial contributions to the conception and design of the work. The research team performed data collection and analysis. All authors contributed to drafting and revising the manuscript, critically reviewed the content, and approved the final version of the manuscript.

## Acknowledgements

The authors would like to thank the nurses who participated in the study. Their time and contribution were invaluable.

## References

1. World Health Organization. State of the World’s Nursing 2020: Investing in education, jobs and leadership. Geneva: WHO; 2020.

2. International Council of Nurses. The Global Nursing Shortage and Nurse Retention. Geneva: ICN; 2021.

3. Buerhaus PI, Auerbach DI, Staiger DO. The recent surge in nurse employment: Causes and implications. Health Aff (Millwood). 2009;28(4):w657–68.

4. Camerino D, Conway PM, van der Heijden BIJM, Estryn-Béhar M, Consonni D, Gould D, et al. Low-perceived work ability, ageing and intention to leave nursing: A comparison among 10 European countries. J Adv Nurs. 2006;55(3):338–48.

5. Ilmarinen J. Work ability—a comprehensive concept for occupational health research and prevention. Scand J Work Environ Health. 2009;35(1):1–5.

6. Trinkoff Am, L. R, Geiger-Brown J, Lipscomb J, Lang G. Longitudinal relationship of work hours, mandatory overtime, and on-call to musculoskeletal problems in nurses. Am J Ind Med. 2006;49(11):964–71.

7. Letvak S. The experience of being an older staff nurse. West J Nurs Res. 2002;24(7):772–90.

8. Shacklock K, Brunetto Y. A model of older workers’ intentions to continue working. Pers Rev. 2011;40(2):252–74.

9. Dall’Ora C, Ball J, Recio-Saucedo A, Griffiths P. Characteristics of shift work and their impact on employee performance and wellbeing: A literature review. Int J Nurs Stud. 2015;52(8):1346–53.

10. Raso R, Fitzpatrick JJ. Older registered nurses and COVID-19. Online J Issues Nurs. 2021;26(2).

11. Holt-Lunstad J, Smith TB, Layton JB. Social relationships and mortality risk: A meta-analytic review. PLoS Med. 2010;7(7):e1000316.

12. Berkman LF, Glass T, Brissette I, Seeman TE. From social integration to health: Durkheim in the new millennium. Soc Sci Med. 2000;51(6):843–57.

13. Victor CR, Bowling A. A longitudinal analysis of loneliness among older people in Great Britain. J Psychol. 2012;146(3):313–31.

14. Simpson R. Presenteeism, power and organizational change: Long hours as a career barrier and the impact on the working lives of women managers. Organization. 1998;5(1):37–62.

15. Cowin LS. Measuring nurses’ self-concept. J Adv Nurs. 2001;33(5):684–92.

16. Elder GH Jr. The life course as developmental theory. Child Dev. 1998;69(1):1–12.

17. Cacioppo JT, Cacioppo S. Social relationships and health: The toxic effects of perceived social isolation. Ann N Y Acad Sci. 2014;1231:17–22.

18. Klinenberg E. Going Solo: The extraordinary rise and surprising appeal of living alone. New York: Penguin Press; 2012.

19. van Manen M. Researching lived experience: Human science for an action sensitive pedagogy. 2nd ed. London: Routledge; 2016.

